# The Trajectory of Depression and Anxiety Among Children and Adolescents over Two Years of the COVID-19 Pandemic

**DOI:** 10.1101/2023.10.04.23296430

**Authors:** Daphne J. Korczak, Ronda F. Lo, Jala Rizeq, Jennifer Crosbie, Alice Charach, Evdokia Anagnostou, Catherine S. Birken, Suneeta Monga, Elizabeth Kelley, Rob Nicolson, Paul D. Arnold, Jonathon L. Maguire, Russell J. Schachar, Stelios Georgiades, Christie L. Burton, Katherine Tombeau Cost

## Abstract

Longitudinal research examining children’s mental health (MH) over the course of the COVID-19 pandemic is scarce. We examined trajectories of depression and anxiety over two pandemic years among children with and without MH disorders. Parents and children 2-18 years completed surveys at seven timepoints (April 2020 to June 2022). Parents completed validated measures of depression and anxiety for children 8-18 years, and validated measures of emotional/behavioural symptoms for children 2-7 years old; children ≥10 years completed validated measures of depression and anxiety. Latent growth curve analysis determined depression and anxiety trajectories, accounting for demographics, child and parent MH. Data were available on 1315 unique children (1259 parent-reports; 550 child-reports). Trajectories were stable across the study period, however individual variation in trajectories was statistically significant. Of included covariates, only initial symptom level predicted symptom trajectories. Among participants with pre-COVID data, a significant increase in depression symptoms relative to pre-pandemic levels was observed. Children and adolescents experienced elevated and sustained levels of depression and anxiety during the two-year period. Findings have direct policy implications in the prioritization and of maintenance of educational, recreational, and social activities with added MH supports in the face of future events.

## 1. Introduction

A large body of research now confirms the negative impact of the onset of the COVID-19 pandemic on children’s mental health (MH) and wellbeing. Meta-analyses of cross-sectional studies examining the effect of the first periods of quarantine on the MH of 22,996 children in Asia and Europe (Panda et al., 2021) and 80,879 children globally (Racine et al., 2021) found that the majority (79%) of children experienced deterioration of their MH or behaviour (Panda et al., 2021) with clinically significant depression and anxiety experienced by 25% and 20% of youth, respectively, during 2020 and early 2021(Racine et al., 2021).

Beyond these cross-sectional data, research examining the longer-term impact of COVID-19 is needed to inform public health, clinical services, and educational planning. Longitudinal studies have shown that children and adolescents experienced increased depression symptoms during the COVID-19 pandemic compared with pre-COVID data (Barendse et al., 2023; Bignardi et al., 2020), with less consistent findings regarding anxiety, of which symptoms in some (Barendse et al., 2023; Bignardi et al., 2020) but not all (Toseeb & Asbury, 2023) studies appeared stable across pandemic timepoints. Indeed, a recent meta-analysis synthesizing changes in depression and anxiety symptoms in 53 studies (40,807 children) across 12 countries found that compared with pre-COVID data, children and adolescents experienced a 26% mean increase in depression symptoms and a 10% mean increase in anxiety symptoms (Madigan et al., 2023). However, only three included studies reported data from 2021, with the remainder reporting outcomes from 2020. This drop-off in longitudinal data is in keeping with the findings of another meta-analysis, in which out of 162 studies that examined during-pandemic children’s mental health, 135 collected data in 2020, 26 collected data in 2021, and one collected data in 2022 (Deng et al., 2023). Thus, in keeping with a recent call to action (Wade et al., 2023), studies that examine the trajectory of children’s mental health outcomes beyond the first year of the pandemic year are needed to determine whether these effects have been maintained, worsened, or have resolved over time.

Pandemic research to date has suggested an increase in mental health impacts for vulnerable children, such as those from socially and economically marginalized groups (Parenteau et al., 2022). Studies conducted early in the pandemic have also suggested that pre-existing health risks, including a history of childhood trauma (Guo et al., 2022), or mental health disorder (Rizeq et al., 2021), may similarly increase vulnerability to the mental health effects of the COVID pandemic. In a cross-sectional survey of 213 parents of children with ADHD in Australia, Sciberras et al. (2022) reported that children with ADHD experienced increased sad or depressed mood, loneliness, and decreased enjoyment in activities (ORs 1.8-6.5, 95% CIs 1.2-10.4) compared to pre-pandemic (Sciberras et al., 2022). In one of the few longitudinal studies examining the mental health of children with a pre-existing MH/NDD condition (collectively referred to herein as MH conditions) from March to October 2020, Toseeb et al (2023) noted that children with ASD experienced an increase in anxiety symptoms that remained elevated over the study period, compared with a general decrease in anxiety symptoms among children with other special education needs and disabilities (Toseeb & Asbury, 2023). To our knowledge, whether children with other pre-existing MH conditions (for example, those with depression or anxiety disorders) similarly demonstrate persistent MH symptom elevations, beyond 2020, is unknown. These data are essential in directing programs and services that are targeted to those most in need.

Thus, the objectives of the current study are (i) to determine the MH effects of the pandemic over time (ii) to determine the MH effects of the pandemic for children with and without pre-existing MH conditions; and (iii) to explore potential risk factors for increased MH symptoms that could serve as targets for preventive intervention. We hypothesized that MH symptoms would be maintained over the two year study period as the disruption to children’s lives (eg school closures) persisted over this time. We further hypothesized that effects would be greater for those with a history of a pre-existing MH condition compared with previously healthy children.

## 2. Methods

### 2.1 Design

Data were collected across seven timepoints that correspond to public health restrictions in the province of Ontario, the most populous province in Canada. Ontario experienced the first wave of the COVID-19 pandemic from March to June 2020. Over the ensuing two years, repeated mass school closures led to the loss of more than 29 weeks of in-person learning for kindergarten to grade 12 students in Ontario, significantly more than the average time lost in Europe (10 weeks) or globally (14 weeks) (UNESCO., 2022). Cancellation/closure of extracurricular activities and recreation centres occurred more variably (Supplemental Figure S1).

### 2.2 Participants and Procedures

Data were collected as part of a large COVID-19 collaboration of four child health and mental health cohorts in Ontario, Canada, each with an existing participant base, to examine the effects of the COVID-19 pandemic on children, youth, and families from the general population and those with pre-existing MH/NDD conditions. Details regarding individual cohorts and the overall study procedure has been previously described (*Citation withheld*). Briefly, the collaboration is comprised of two clinically-referred MH and neurodevelopmental cohorts and two community cohorts: (i) Mental Health: children/adolescents referred to an outpatient MH clinic for evaluation of MH concerns including, but not limited to, depression and anxiety disorders, attention-deficit/hyperactivity disorder (ADHD), obsessive-compulsive disorder (OCD), disruptive behaviour disorders; (ii) NDD: children/adolescents in the community with neurodevelopmental disorders, including autism spectrum disorders (ASD), ADHD, OCD, and intellectual disability (*Citation withheld*); (iii) Primary Care: healthy children recruited from birth to age five years and participating in a primary care practice-based research network (*Citation withheld*); and (iv) Population-based: a population-based sample of children/adolescents recruited at an urban science museum (*Citation withheld*). These were selected as established cohorts with a diverse, existing participant base that include children birth to 18 years enhanced for children and adolescents with pre-existing MH and NDDs.

Parents who had previously consented to be contacted were sent an invitation email and link for their child’s attention or to send to their child’s email if their child was aged 10 to 18 years and interested in participating. The email contained all information pertinent for consent to participation, including purpose of the study, risks and benefits of participation, and confidentiality. Participants were not selected for invitation based on any specific characteristics. Parents and children completed online surveys via the survey application REDcap (Harris et al., 2009) in April 2020 and at regular intervals at follow up timepoints between April 2020 and June 2022. All procedures were the same for all cohorts and were approved at all institutional research ethics boards. Written informed consent was obtained from all participants.

Data were divided into seven time points based on time of collection: April to June 2020 (T1), July to August 2020 (T2), September to November 2020 (T3), December 2020 to May 2021 (T4), June to October 2021 (T5), November 2021 to March 2022 (T6), April to June 2022 (T7). These time points were created to examine MH outcomes during periods that correspond to both important seasonal influences in children’s lives (e.g., summer holiday) as well as the experience of the COVID-19 pandemic and related lockdowns in Ontario.

### 2.3 Measures

#### 2.3.1 Mental Health

Child and adolescent MH outcomes were assessed at each study timepoint. Depressive symptoms (children ages 8-18 years) were measured using the Major Depressive Disorder subscale of the parent-reported Revised Child Anxiety and Depression Scales-Parent Version (RCADS-P) (Ebesutani et al., 2015). The 10-item scale converts to standardized T-scores, where a T-score of 65 indicates borderline clinical range, in the top 7% of children in the same grade and sex. Child-report of depression (children 10-18 years) was determined using the 20-item Centre for Epidemiological Studies Depression Scale for Children (CES-DC) (Fendrich et al., 1990). Items are rated on a scale of 0 to 3 and assess depressive symptoms over the past week. Total sum scores range from 0 to 60, with higher scores indicate greater severity of depression. Scores of 15 or higher indicate a positive screen for clinically significant depressive symptoms(Fendrich et al., 1990).

Anxiety symptoms were measured by parent-report (8-18 years) and child-report (10-18 years), using the 9-item Generalized Anxiety Disorder subscale of the Screen for Child Anxiety Related Disorders (SCARED) instrument (Birmaher et al., 1997). Scores of 9 or higher indicate a positive screen for clinically significant anxiety symptoms. Parents of children ages 2 to 7 years old completed either the Preschool Strengths and Difficulties Questionnaire (SDQ) (2-4 years) or the SDQ (Goodman, 1997) (5-7 years) to report on their child’s MH problems. In population-based samples, total difficulties scores greater than the 90^th^ percentile have been suggested to indicate clinical significance (Koskelainen et al., 2000).

#### 2.3.2 Predictor Variables

Predictor variables were assessed at pandemic onset (T1).

##### Parent Anxiety

Parent anxiety was measured at Time 1 using the 7-item Generalized Anxiety Disorder assessment (GAD-7) (Spitzer et al., 2006), a widely used and valid instrument with increasing scores indicating increasing anxiety.

##### Demographics

Parent-reported annual household income in Canadian dollars, child age, race/ethnicity, sex, and pre-COVID MH/NDD history were determined at T1 using items adapted from the CRISIS questionnaire (Nikolaidis et al., 2021), an instrument designed by an international collaboration to examine MH during the COVID-19 pandemic.

### 2.4 Data Analysis

One measurement per participant was retained within each time point based on data completion, such that for participants with more than one data per timepoint, the data point with the most complete data was selected for inclusion. If there was no difference in data completion between data points, the most recent data point was selected. Participants with data on MH outcomes across two or more time points were included. In the event of participant siblings, only one sibling per family was retained in the dataset, with the sibling with the most complete data selected for retention. In the event that sibling data was equally complete, the eldest sibling was selected.

Data were analysed in RStudio version 1.3.1093 (R Team, 2020). Descriptive statistics for all variables were calculated. To determine the MH effects of the pandemic over time, linear growth curve models were estimated across the seven time points. First, a set of unconditional linear growth curve models were estimated to create models with the best fit to the data based on the fit indices (Duncan & Duncan, 2004). Unconditional growth curve models were estimated for each MH outcome (RCADS-P, CES-DC, SCARED-P, SCARED-C, SDQ) and stratification (MH vulnerability: MH history, no MH history) separately, using the RStudio *lavaan* package,(R Team, 2020) using full information maximum likelihood estimator, which allows all available data to be used when there are missing data.

Standardized root mean square residual (SRMR), root mean square error of approximation (RMSEA), comparative-fit index (CFI), and Tucker-Lewis index (TLI) were used to evaluate model fit indices against rigorous standards (Hu & Bentler, 1999). Pre-pandemic depression [RCADS-P] data were available for a subset of the participants (n = 330), collected from October 2018 to March 2020. To further understand the MH effects of the pandemic compared with pre-pandemic MH symptom levels, we compared pre-pandemic with T1 RCADS-P scores using paired sample *t*-tests. To determine whether sociodemographic characteristics for participants with pre-pandemic depression score were different from those without pre-pandemic depression scores, we compared these groups using chi-square tests of equal proportions or Welch’s *t*-tests (Supplemental Table S1). To determine whether RCADS-P scores during the pandemic were different for individuals with pre-pandemic data, a two-way mixed analysis of variance (ANOVA) was conducted to examine differences in depression scores at each timepoint, between participants with and without pre-pandemic depression scores. To examine whether symptom trajectories over time had potential clinical implications, the proportion of children above the clinically significant threshold for depression or anxiety at different study timepoints was compared using McNemar χ^2^ tests for paired data.

To understand the MH effect over time for children with and without pre-existing MH conditions, and to explore other potential risk factors for increased MH symptoms that could serve as targets for preventive intervention, a set of conditional models with time invariant covariates were examined. Conditional models for pre-existing MH diagnosis included ethnicity, income, sex, age, and parental anxiety at Time 1 and assessed whether the covariates were statistically significantly associated with inter-individual differences in either the initial level of MH symptoms (i.e., the intercept at T1) or in MH symptom change over time (i.e., the slope from T1 through T7 inclusive). When including exogenous covariates in the conditional models, we specified incomplete data for the exogenous covariates be incorporated in the model estimation. Conditional models stratified by MH vulnerability were estimated separately for parent and child report of depression (RCADS-P_noMH_, RCADS-P_MH_, CES-DC_noMH_, CES-DC_MH_) and for parent and child report of anxiety (SCARED-P_noMH_, SCARED-P_MH_, SCARED-C_noMH_, SCARED-C_MH)_.

## 3. Results

Of the 1,681 children with data on any outcome measure, completed measures for at least two timepoints were available on 1,315 unique children ages 2 to 18 years (1259 parent-report and 550 child/youth-report; response rate 78%). After excluding siblings, the analytic sample comprised 988 unique children ages 8-18 years, 932 with parent-reported outcomes (56.2% male; mean age 11.79 [SD 2.9] years) and 550 with child-report outcomes (52.9% male; mean age 12.9 [SD 2.5] years). The analytic sample among 2-to 7-year old children comprised 327 unique children (50.8% male; mean age 4.37 [SD 1.37] years). Sociodemographic characteristics are described in Table 1.

**Table 1.** Demographic Information.

|  | Parent Report<br>(8-18 years) | Child (Self) Report<br>(10-18 years) | Parent Report<br>(2-7 years) |
| --- | --- | --- | --- |
|  | *n* = 932 | *n* = 550 | *n* = 327 |
| Child age – Mean (*SD*) | 11.79 (2.90) | 12.91 (2.48) | 4.37 (1.37) |
| Assigned sex of child – % (*n*) |  |  |  |
| Male | 56.2% (524) | 52.9% (291) | 50.8% (166) |
| Female | 43.7% (407) | 46.7% (257) | 48.6% (159) |
| Other | 0.1% (1) | 0.2% (1) | 0% (0) |
| Did not respond | 0% (0) | 0.2% (1) | 0.6% (2) |
| Parent-reported Family Income – % (*n*) |  |  |  |
| Less than \$80,000 | 23.3% (217) | 21.3% (117) | 14.7% (48) |
| Over \$80,000 | 61.8% (576) | 57.6% (317) | 75.5% (247) |
| Did not respond | 14.9% (139) | 21.1% (116) | 9.8% (32) |
| Ethnicity/Ancestry – % (*n*) |  |  |  |
| European (non-Indigenous North American) | 66.0% (615) | 60.5% (333) | 48.0% (157) |
| Non-European (single ancestry origin) | 16.7% (156) | 17.3% (95) | 24.8% (81) |
| Multiple ancestry origin | 15.1% (141) | 16.2% (89) | 10.7% (35) |
| Did not respond | 2.1% (20) | 6.0% (33) | 16.5% (54) |
| Prior Mental Health Diagnoses* – % (*n*) | 68.5% (638) | 66.2% (364) | 4% (13) |

\*As assessed at timepoint 1 (T1), pandemic onset (April-June 2020).

### Trajectories of Depression During the Pandemic (April 2020 – June 2022)

#### Model fit indices

Model fit indices for all linear unconditional and conditional models examining depression outcomes were excellent (Supplemental Table S2). Excellent fit indices to the linear models indicated that the average trajectories assumed a linear shape over the course of the pandemic for each of the depression outcomes (SDQ, RCADS-P, CES-DC) by each of the stratifications (noMH, MH).

#### Conditional models of depression: MH effects over time

For children ages 8-18 years, the means of the slope of the trajectories, and nearly all predictors of the slope, were statistically non-significant for all models (Table 2, Supplemental Tables S3, S4), indicating that on average, symptom levels did not change over the study period, irrespective of informant or MH history (Figure 1). However, all depression models indicated statistically significant variance in the slopes, regardless of informant, indicating that there were inter-individual differences among children in the amount and direction of change in depression symptoms over the course of all seven timepoints (Supplemental Tables S3, S4). This variance in the trajectories was not explained by included covariates for nearly all models, indicating that individual variation in change over the course of the pandemic was related to factors outside of those included here.

**Figure 1A, B.**
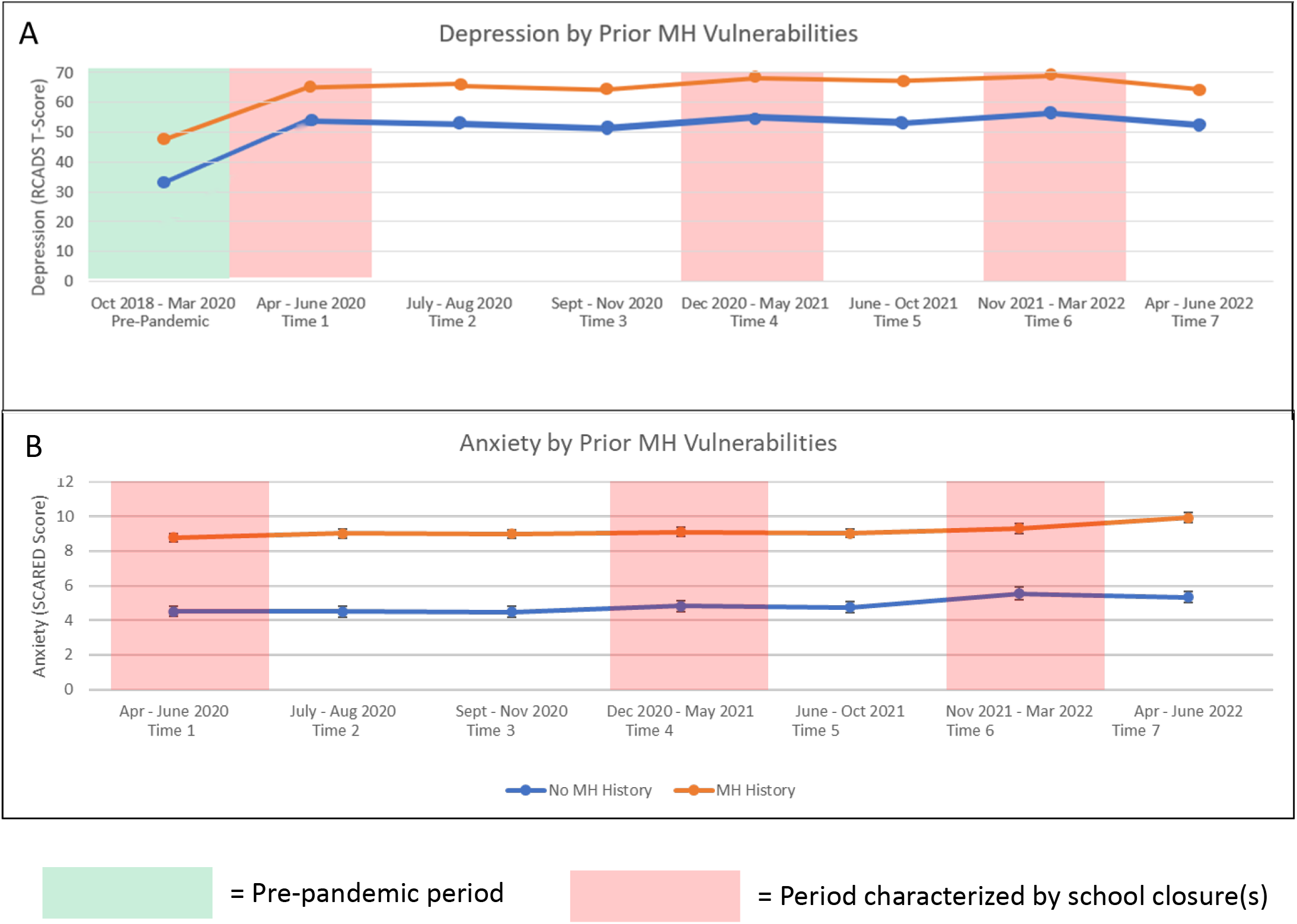
Parent-Report Trajectories of Depression and Anxiety. *Note*. Error bars indicate standard errors of the mean. MH = Prior mental health history. Depression is measured with RCADS (Revised Children's Anxiety and Depression Scale). Anxiety is measured with SCARED (Screen for Child Anxiety Related Disorders). Figure 1A pre-pandemic sample sizes (green shaded columns): 8-12 years n = 230, 13-18 years n = 100. Figure 1A pandemic sample sizes (pink shaded and unshaded columns): no MH history n = 293, MH history *n* = 635. Figure 1B sample sizes: no MH history *n* = 289, MH history *n* = 616.

**Figure 1C,D.**
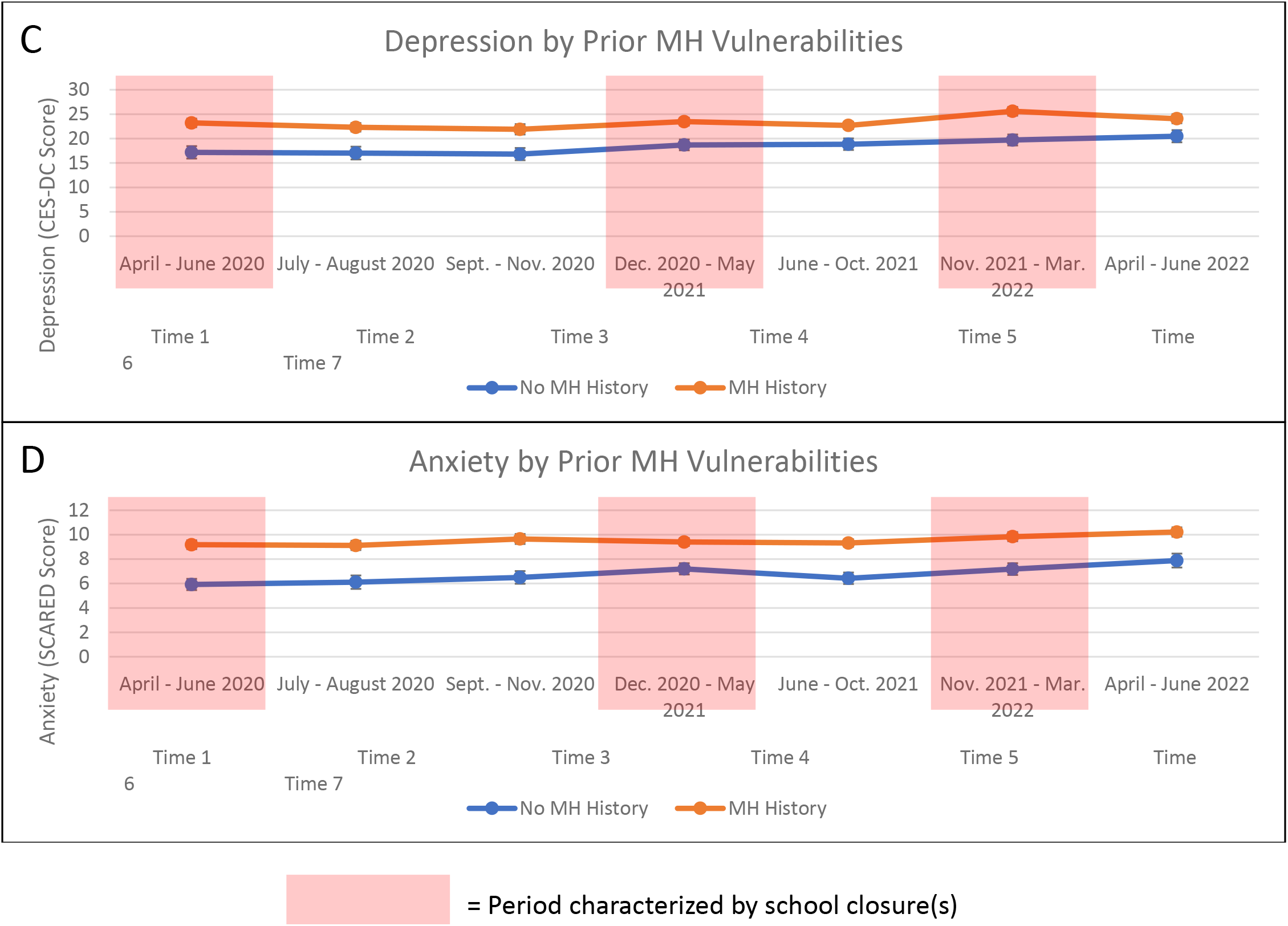
Child-Report Trajectories of Depression and Anxiety. *Note*. Error bars indicate standard errors of the mean. MH = mental health. Depressions is measured with CES-DC (Center for Epidemiologic Studies Depression Scale for Children). Anxiety is measured with SCARED (Screen for Child Anxiety Related Disorders). Figure 1C sample sizes: no MH history *n* = 186, MH history *n* = 363. Figure 1D sample sizes: no MH history *n* = 186, MH history *n* = 364.

**Figure 1E.**
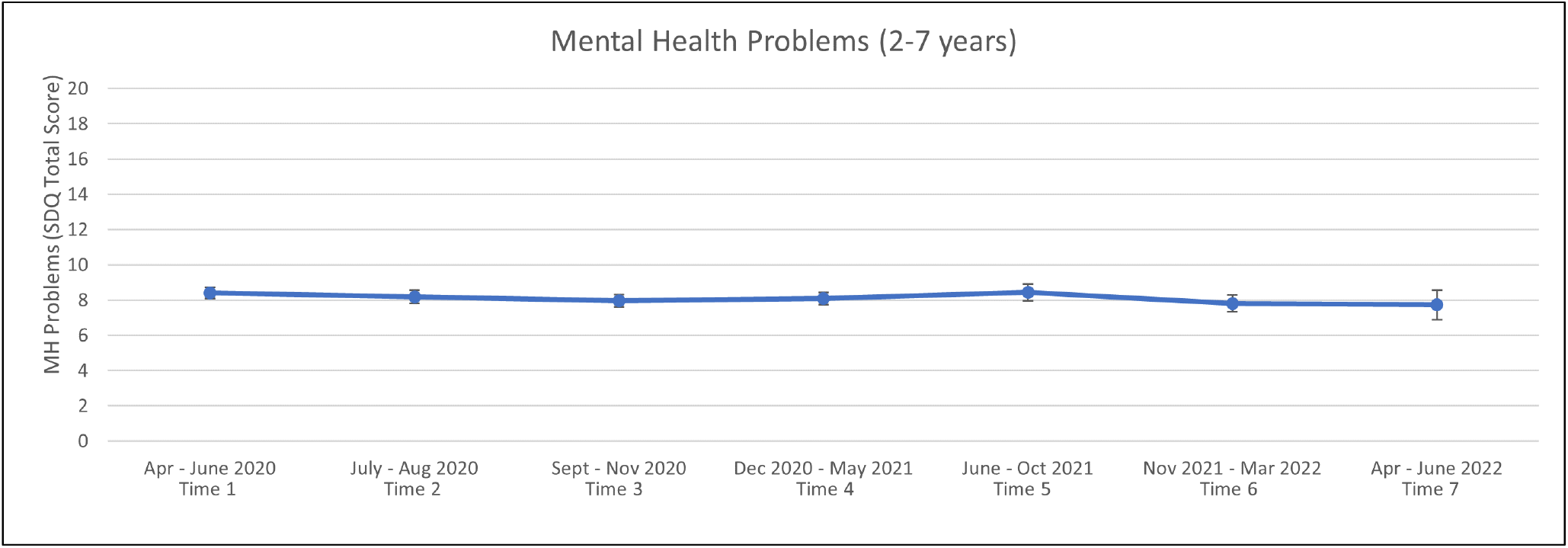
Parent-Report Trajectories of Mental Health Problems. *Note*. Error bars indicate standard errors of the mean. Mental health problems is measured with the Strengths and Difficulties Questionnaire (SDQ). Sample size: *n* = 327

**Table 2.** Conditional Model Parameters (All Covariates) For Children and Youth Ages 8-18.

|  |  | RCADS-P |  | SCARED-P |  | CES-DC |  | SCARED-C |  |
| --- | --- | --- | --- | --- | --- | --- | --- | --- | --- |
| Diagnostic Group | Covariates |  |  |  |  |  |  |  |  |
| No MH | Intercept | **47.03 (4.04)** | < .001 | 1.45 (1.06) | .17 | -0.68 (5.33) | .90 | -0.44 (2.24) | .84 |
|  | Variance of Intercept | **120.08 (19.21)** | < .001 | **11.03 (1.48)** | < .001 | **116.40 (17.57)** | < .001 | **16.13 (2.65)** | < .001 |
|  | Income | **-5.96 (2.04)** | .003 | -0.70 (0.61) | .25 | -3.90 (2.56) | .13 | -1.83 (1.03) | .08 |
|  | Ethnicity | 2.95 (2.16) | .17 | 0.25 (0.66) | .70 | -1.46 (2.38) | .54 | -0.64 (0.98) | .52 |
|  | Sex | -1.64 (1.55) | .29 | **0.98 (0.46)** | .03 | 3.17 (1.81) | .08 | 0.76 (0.72) | .29 |
|  | Age | 0.28 (0.29) | .34 | 0.12 (0.08) | .14 | **1.17 (0.45)** | .01 | **0.47 (0.17)** | .01 |
|  | Parent Anxiety | **1.42 (0.23)** | < .001 | **0.29 (0.05)** | < .001 | **1.04 (0.23)** | < .001 | **0.39 (0.10)** | < .001 |
|  | Slope | 0.11 (0.81) | .90 | **0.46 (0.22)** | .04 | -1.13 (1.02) | .26 | -0.40 (0.42) | .34 |
|  | Variance of Slope | **2.39 (0.78)** | .002 | **0.21 (0.05)** | < .001 | **3.03 (0.86)** | < .001 | **0.39 (0.13)** | .003 |
|  | Income | -0.004 (0.45) | .99 | -0.19 (0.11) | .09 | 0.34 (0.45) | .37 | -0.03 (0.88) | .88 |
|  | Ethnicity | -0.50 (0.42) | .324 | -0.14 (0.13) | .29 | -0.02 (0.45) | .97 | 0.16 (0.22) | .46 |
|  | Sex | **0.66 (0.32)** | .04 | 0.16 (0.09) | .07 | 0.64 (0.37) | .09 | **0.36 (0.15)** | .02 |
|  | Age | 0.01 (0.06) | .94 | -0.02 (0.02) | .29 | 0.11 (0.08) | .17 | 0.04 (0.03) | .18 |
|  | Parent Anxiety | -0.06 (0.05) | .23 | -0.00 (0.01) | .97 | -0.05 (0.05) | .35 | -0.01 (0.02) | .55 |
|  | Intercept-Slope Correlation | **-8.04 (3.29)** | .01 | -0.37 (0.20) | .06 | **-9.09 (3.20)** | .005 | -0.52 (0.48) | .28 |
| Diagnostic Group | Covariates |  |  |  |  |  |  |  |  |
| MH History | Intercept | **56.46 (3.21)** | < .001 | **4.44 (0.95)** | < .001 | 4.64 (4.07) | .25 | 0.57 (1.61) | .72 |
|  | Variance of Intercept | **181.97 (15.13)** | < .001 | **18.96 (1.20)** | < .001 | **121.70 (11.39)** | < .001 | **19.63 (1.63)** | < .001 |
|  | Income | -0.88 (1.54) | .57 | -0.12 (0.47) | .81 | -2.53 (1.68) | .13 | -0.88 (0.70) | .21 |
|  | Ethnicity | 0.95 (1.80) | .60 | -0.33 (0.54) | .54 | 1.48 (1.87) | .43 | -0.18 (0.77) | .82 |
|  | Sex | 1.38 (1.29) | .28 | **1.00 (0.40)** | .01 | **4.15 (1.44)** | .004 | **2.21 (0.56)** | < .001 |
|  | Age | 0.12 (0.22) | .58 | **0.16 (0.07)** | .01 | **1.13 (0.28)** | < .001 | **0.54 (0.11)** | < .001 |
|  | Parent Anxiety | **0.97 (0.13)** | < .001 | **0.28 (0.04)** | < .001 | **0.33 (0.13)** | .01 | **0.13 (0.05)** | .01 |
|  | **Slope** | -0.38 (0.64) | .55 | 0.16 (0.17) | .36 | 0.88 (0.84) | .29 | 0.26 (0.34) | .45 |
|  | **Variance of Slope** | **3.35 (0.66)** | < .001 | **0.27 (0.06)** | < .001 | **3.24 (0.68)** | < .001 | **0.52 (0.09)** | < .001 |
|  | Income | -0.09 (0.31) | .78 | 0.13 (0.08) | .13 | -0.12 (0.36) | .74 | -0.03 (0.14) | .83 |
|  | Ethnicity | 0.37 (0.37) | .32 | 0.16 (0.10) | .12 | -0.11 (0.36) | .76 | -0.15 (0.15) | .33 |
|  | Sex | 0.21 (0.26) | .43 | 0.11 (0.08) | .16 | 0.002 (1.00) | .99 | 0.01 (0.12) | .97 |
|  | Age | 0.03 (0.05) | .48 | -0.01 (0.01) | .41 | -1.03 (0.06) | .30 | -0.01 (0.02) | .59 |
|  | Parent Anxiety | 0.003 (0.02) | .90 | -0.01 (0.01) | .21 | 0.05 (0.03) | .16 | 0.01 (0.01) | .23 |
|  | **Intercept-Slope Correlation** | **-6.45 (2.53)** | .01 | **-0.53 (0.22)** | .01 | **-10.52 (2.31)** | < .001 | **-1.52 (0.33)** | < .001 |

*Note:* Estimates are in unstandardized units. Bolded estimates are statistically significant at $p < .05$ . Standard error of the means are reported in parentheses. No MH= No prior mental health history; MH History = Prior mental health history. Depression is measured with parent-reported RCADS-P (Revised Children's Anxiety and Depression Scale) and child reported CESDC (Center for Epidemiological Studies Depression Scale for Children). Anxiety is measured with parent and child- reported SCARED (Screen for Child Anxiety Related Disorders; SCARED-P, SCARED-C). \*Child-report of depression/anxiety (CES-DC/SCARED-C) collected from children age 10 years and older

For 2 to 7 year old children, the slope of the trajectory of mental health problems was non-significant and indicated a stable level of emotional and behavioural difficulties of children in this age group over the study period (Figure 1E). The variance on the slope was significant, indicating individual differences in trajectories across the pandemic (Supplemental Table S5). However, none of the predictors we included explained the inter-individual differences in change over time in this age group (Supplemental Table 5).

#### Depression models: Effect of MH history

Children ages 8 to 18 years MH conditions experienced greater depression symptoms compared with previously healthy children at onset of the pandemic (T1; the intercept), irrespective of informant (RCADS-P 65.34 [SD 17.09] vs. 53.61 [SD 15.54], *p* < .001; CES-DC 23.21 [SD 13.07] vs 17.18 [12.78], p<.001; Supplemental Table 6). There was also statistically significant variance in the intercepts, indicating inter-individual differences in the levels of depression symptoms at the start of the pandemic across all informant and MH history groups (Table 2; all p’s<.001). Increased parental anxiety was significantly associated with higher child depression at the start of the pandemic (T1, ie, the intercept) for depression regardless of informant or MH vulnerability (*p*-values <= .01; Table 2). This suggests that children whose parents were experiencing higher levels of anxiety had higher depression symptoms at the start of the pandemic as compared with children whose parents had lower levels of anxiety.

Among 2–7-year-old children, very few children had a MH history (n = 13 [4%]), therefore all children were included in one analysis without stratification by MH vulnerability.

### Changes in Depression Symptoms From Pre-Pandemic to Pandemic Onset (T1)

Among children for whom pre-COVID data were available, depression symptoms increased significantly (RCADS-P scores) following the onset of the COVID pandemic (pre-COVID 28.01 [SD 15.48] vs during-COVID: 61.33 [SD 17.33], *p* < .001; Supplemental Table S1). The trajectories of participants with pre-COVID depression scores were comparable with those without pre-COVID data across all timepoints (T1-T7) (*F*(6, 1020) = 0·61, p = 0·72, *η_p_^2^* = 0·004; Supplemental Table S1). Compared with participants without pre-pandemic data, participants with pre-COVID depression data were slightly younger (mean 11.43 vs 11.98 years, *p*=.004), with no differences in participant sex, race/ethnicity, or family income (Supplemental Table S1).

### Clinically Significant Depression Symptoms

Figure 2 details the proportion of children that met or exceeded clinically significant thresholds for depression symptoms at each timepoint by MH history. Among previously healthy children, the proportion experiencing depression symptoms in the clinically significant range increased from 49.1% at T1 to 60.3% at T6 (*p*=.03; Figure 2). Among children with MH conditions, the proportion of children experiencing symptoms in the clinically significant range for depression increased from 68.2% at T1 to 77.5% at T6 (*p*=.003; Figure 2, Supplemental Table S7). Depression symptoms by age across study time points indicated that potentially clinically significant depression symptoms were greater among older (13-18 years) versus younger (8-12 years) children (Supplemental Table S6, Supplemental Figure 2).

**Figure 2.**
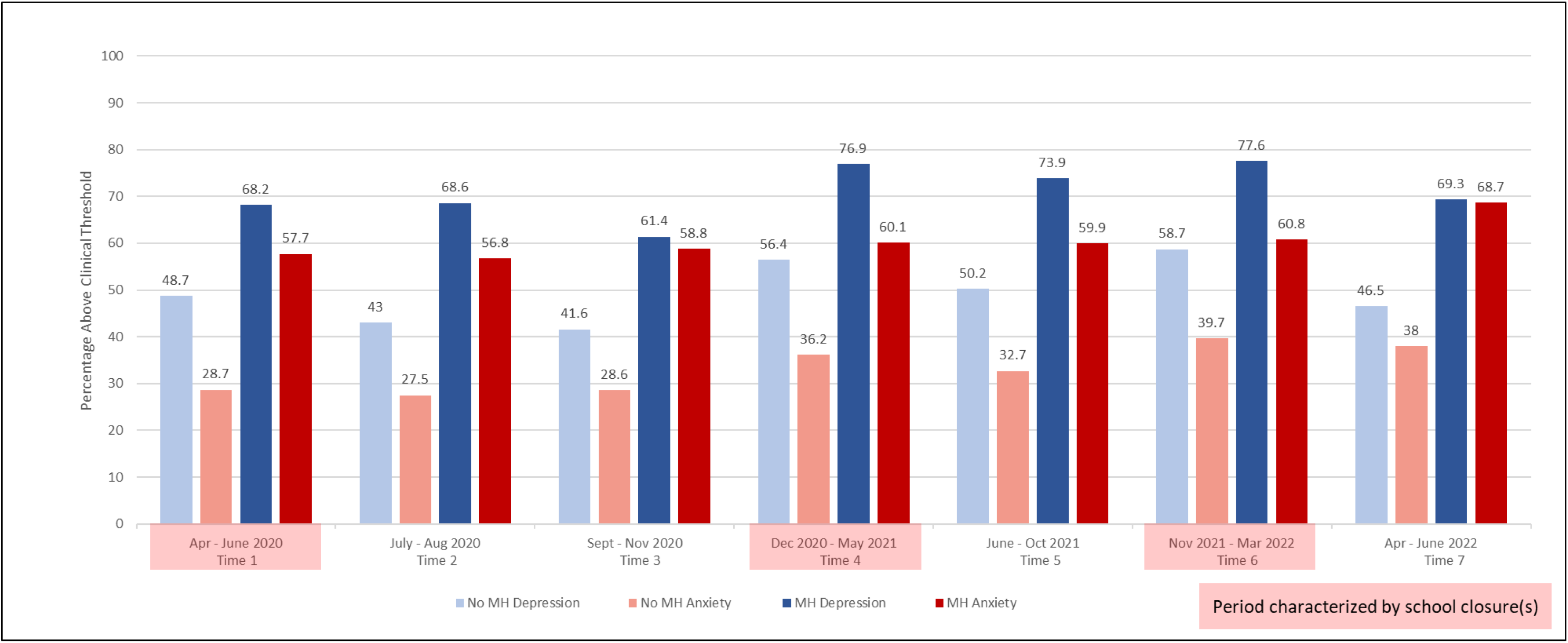
Clinically Significant Depression or Anxiety Symptoms Among Children With and Without a Pre-COVID MH History. *Note*. Bar chart displays percentage of children who meet or exceed clinical threshold for Depression (RCADS or CES-DC reported by parents and children, respectively) and Anxiety (SCARED). Missing data were removed prior to visualization.

### Trajectories of Anxiety During the Pandemic (April 2020-June 2022)

#### Model fit indices

Model fit indices for all linear unconditional and conditional models examining anxiety outcomes were excellent (Supplemental Table S2). Excellent fit indices to the linear models indicated that the average trajectories assumed a linear shape over the course of the pandemic for each of the anxiety outcomes (SCARED-P, SCARED-C) by each of the stratifications (noMH, MH).

#### Conditional models of Anxiety: Effects over time

For children ages 8-18 years, the means of the slopes of the trajectories were not statistically significantly different from zero in almost all models, indicating that, on average, children’s level of anxiety symptoms were stable over the study period (Table 2) irrespective of informant or age, with the single exception of increasing anxiety in previously healthy children as reported by parents but not by children (Figures 1A-D; Table 2). However, all anxiety models indicated statistically significant variance in the slopes, regardless of age, informant, or MH history, indicating that there were inter-individual differences among children in the amount and direction of change in anxiety symptoms over the course of all seven timepoints (Table 2, Supplemental Tables S3, S4). With a single exception (females_noMH_ self-reported increasing anxiety symptoms over time), this variance in the trajectories was not explained by included covariates (Table 2), indicating that individual variation in change over the course of the pandemic was related to factors outside of those included here.

#### Effect of MH History

In the MH stratified models, the mean of the intercept for parent-reported (SCARED-P) but not child-reported (SCARED-C) anxiety in the MH group was statistically significant, indicating that children’s anxiety symptoms was significantly higher than zero at the start of the pandemic for children with a pre-existing MH condition. Children with MH conditions had higher anxiety scores at the onset of the pandemic (T1) compared with previously healthy children regardless of informant (SCARED-P_MH_: 8.77 [SD 5.16] vs. SCARED-P_noMH_: 4.51 [SD 4.17], *p* < .001; SCARED-C_MH_: 9.18 [5.36] vs. SCARED-C_noMH_: 5.92 [SD 4.69], *p* < .001; Supplemental Table 6).

There was significant variance in the intercepts, indicating the different children had different levels of anxiety symptoms at the start of the pandemic across informants and MH vulnerability groups (all p’s < .001; Table 2). While there are some differences in the predictors of the intercepts across models, increased parental anxiety was significantly associated with higher child anxiety at the start of the pandemic (*p*s <= .01; Table 2). This indicates that children whose parents were experiencing higher levels of anxiety had higher anxiety symptoms at the start of the pandemic as compared with children whose parents had lower levels of anxiety. Anxiety symptoms by MH history across study time points are presented in Table 2.

#### Clinically Significant Anxiety Symptoms

Among previously healthy children, the proportion experiencing anxiety symptoms in the clinically significant range increased significantly from 28.7% at T1 to 40% at T6, (*p*=.04) its highest point. Among children with MH conditions, the proportion of children experiencing anxiety symptoms in the clinically significant range increased from 57.7% at T1 to a high of 68.7% at T7 (*p*=.004, Figure 2B, Supplemental Table S7). Clinically significant anxiety symptoms across age groups indicated that potentially clinically significant anxiety symptoms were greater among older (13-18 years) versus younger (8-12 years) children (Supplemental Table S6, Supplemental Figure 2).

## 4. Discussion

This study finds that children experienced elevated levels of depression and anxiety symptoms over the two years following the onset of the COVID-19 pandemic in Canada. Increased rates of depression and anxiety were observed among school aged children regardless of MH history, however, those with prior MH conditions experienced the greatest symptom burden overall. No evidence of adaptation to pandemic-related restrictions was observed despite wide fluctuations in community viral transmission of COVID-19 over the same time period. Further, among participants with pre-COVID data on depression, these scores represent a significant increase in depression symptoms relative to pre-pandemic levels.

These results confirm and extend cross-sectional (Racine et al., 2021) and longitudinal (Thorisdottir et al., 2021) reports demonstrating significant and increased MH symptoms during the pandemic. In general, higher MH symptoms were experienced by older children and by those with a pre-existing MH disorder compared with children who were younger or previously healthy at the onset of the pandemic. The magnitude of depression and anxiety symptoms that children experienced during the COVID-19 pandemic in our study is also higher than those reported prior to the pandemic in previous research. With respect to depression, children and adolescents in our study experienced significantly higher symptom scores during the COVID-19 pandemic than those reported prior to the pandemic, using the same measures (RCADS-P, CES-DC) across mental health history groups (all ps < .001) (Breaux et al., 2021; Duttweiler et al., 2022; Feurer et al., 2022; Walters et al., 2021). With respect to anxiety, children and adolescents in our study with no prior mental health disorder also experienced significantly higher symptom scores during the COVID-19 pandemic than those reported prior to the COVID-19 pandemic, using the same measures (SCARED-P, SCARED-C; all ps <.001).(Adegboye et al., 2021; Hawes et al., 2022).

Our findings underscore the importance of the initial responses (i.e., the intercepts in our models) among children, as this initial response was, on average, maintained over the two-year study period. Therefore, the determinants of children’s initial response are particularly salient. Specifically, only pre-existing MH disorder and parental anxiety were consistent factors associated with children’s initial emotional response to the pandemic, indicating the need for further research examining other factors (e.g., effects of parent MH, family conflict, child MH service provision) as potential targets for intervention.

Within diagnostic groups, however, we found significant individual variation with respect to the trajectory of depression and anxiety symptoms over time. None of the covariates that we measured (age, sex, income, ethnicity, pre-existing MH disorder, or parental anxiety) consistently predicted the course of an individual child’s distress. This suggests that other, unmeasured, factors have impacted the trajectories of children’s depression and anxiety symptoms. Previous research has confirmed the impact of factors that are more proximal to the child then the factors in our models, including social isolation, online learning, increased screen time, decreased physical activity, and material deprivation (Cost et al., 2021; LaForge-MacKenzie et al., 2022; Rizeq et al., 2021; Tsujimoto et al., 2022). Future research examining the impact of these environmental and family factors on children’s mental distress is needed.

We found that children and adolescents experienced a greater magnitude of depression compared with anxiety symptoms during the pandemic. These findings are consistent with research highlighting COVID-related impacts on school, social engagement, and activity restrictions on children’s sadness, sense of purpose, and hopelessness (Tsujimoto et al., 2022; Yard et al., 2021), which can perpetuate depressed mood among youth. Our findings also align with a global trend, and are consistent those of a recent meta-analysis of longitudinal studies examining depression and anxiety symptoms pre-versus during-the COVID-19 pandemic (Madigan et al., 2023), which reported a more significant increase in depression than in anxiety symptoms among children and adolescents early in the pandemic (primarily from 2020). Thus, our findings confirm and extend those of previous research noting the greater reactivity and episodicity of mood compared with anxiety, which is known to demonstrate greater intra-individual stability over time (Hovenkamp-Hermelink et al., 2019).

Over half of previously healthy children and over 70% of children with prior MH problems reported depressive symptoms in the clinically significant range at several study timepoints. This represents a larger proportion of screen-positive children than expected based on pre-COVID research, in which self-report rates of up to 24% have been reported among predominantly older samples of adolescents in primary care settings (Zuckerbrot et al., 2018). The persistence of MH symptoms in the clinically significant range over the 2-year timeframe also highlights the chronic nature of the stressor that children in Ontario, Canada faced, despite fluctuations in community viral transmission or economic re-opening. Children demonstrated some symptom decrease during the months in which public health restrictions were lightened, and in-person school resumed (T3, T7) compared with periods of school shutdown/lockdowns (T1, T4, T6). However, a high degree of symptom burden was retained, suggesting that many children experienced negative impact of the broader pandemic context.

.Children with no previous MH disorder exhibited significant increases in depression symptoms in particular, compared with pre-pandemic estimates of approximately 7-12% (Avenevoli et al., 2015). Depression group means of previously healthy children increased from pre-pandemic non-depressed levels to those hovering at or near clinically significant thresholds for the duration of the pandemic study period. This has significant public health and resource implications, as the majority of children in the population had no pre-pandemic MH disorder. Thus, even small increases in the proportion of previously well children experiencing clinically significant depression and anxiety symptoms will have significant impacts on MH service needs.

Consistent with previous literature (De Los Reyes et al., 2015; van der Ende et al., 2012), children reported greater depression and anxiety symptoms than parents detected, when considering children’s report of average scores in the clinically significant range as well as the magnitude of symptoms above clinical thresholds. This was true across MH history groups, though particularly notable for depressive symptoms among previously healthy children. Among these groups, parents reported children’s symptoms, on average, as being below clinical thresholds, whereas children reported their distress to be clinically significant. Children may have reported their symptoms to be greater than parents detected for several reasons, including reluctance to report sadness, worry or guilt due to difficulty articulating distress, out of fear that their experiences may be minimized, in attempt to protect an already stressed or unwell parent, or due to the inherent pessimism of depression. Decreased parental detection of children’s symptoms among previously considered low risk groups (no prior MH condition) may also be, in part, due to a combination of a low index of suspicion and decreased parental sensitivity among parents experiencing increased stress due to the COVID pandemic (Rizeq et al., 2021).

## Limitations

To our knowledge, this is the first study to examine the longitudinal association of the COVID-19 pandemic on the MH of children with and without a history of psychiatric illness over two years of the pandemic. The current study has several strengths, including participation of a large sample of children, implementation of validated measures of depression and anxiety, availability of pre-COVID data on a subset of the sample, and inclusion of the report of children themselves. However, several limitations are also noted. The under-representation of low-income and racialized individuals limits generalizability of study findings to these populations. Given research reporting a disproportionate impact of the COVID-19 pandemic on marginalized groups (Kirby, 2020), the current study may underestimate the MH impacts for racialized youth and for children in lower income families. Also, we were unable to examine the impact of parental depression or of fluctuations in parent MH over the study period, as parental anxiety and depression were highly collinear in our model, allowing the inclusion of only one of these variables. Given the significance of parental anxiety on children’s initial responses in the current study, however, and as parent MH is modifiable, future studies should examine the interactional association of parent and child MH over time.

## Conclusions

We observe that children and adolescents experienced elevated levels of MH symptoms during the pandemic which have been sustained across the two-year pandemic study period. Symptom trajectories varied at the level of the individual; however, individual variations were not accounted for by demographic data, personal history of mental disorder, or parental anxiety. Findings suggest an urgent need to prioritize children in considering policy, educational, and health-implications of public health restrictions and support the importance of social interactions and activities in children’s lives. Results further suggest that increased MH supports for children and parents will be needed during the transition to a post-pandemic recovery.

## Data Availability

Data produced in the present work are contained in the manuscript.

## Declarations

## Acknowledgements

The Ontario COVID & Kids Mental Health Collaboration acknowledges the families, children, and youth who have generously contributed their time and their experience participating in this research. The research team also acknowledges the trainees, analysts, project coordinators, and cohort staff whose dedication has made this research possible.

## Availability of Data and Code

The data that support the findings of this study are not openly available and are available from the corresponding author upon reasonable request (including a study outline), subject to review. Data analytic plans and full statistical reports are available in the supplemental material.

## Author Contribution

DK, with input from JC, RFL, EA, KTC, AC, CSB conceived the research question and study design. DK supervised all aspects of the study, and wrote the first draft of the manuscript. KTC contributed to and supervised all analyses, contributed to the conceptualization of the study. JR and RFL ran the analyses, contributed to, and reviewed drafts of the manuscript. RFL prepared and presented results and appendices, including visualization of data. All authors contributed to the selection and development of study instruments, data collection, and study administration. JC, EA, SM, AC and CSB contributed to the conceptualization and study design and reviewed and edited drafts of the manuscript. EK, RN, JM, PA, RS, SG, and CLB, reviewed and edited drafts of the manuscript. All authors approved the final draft. All authors had full access to all data in the study and had final responsibility for the decision to submit for publication.

## Ethics Approval

The study was approved by the institutional research ethics board at the lead research site (SickKids, 1000070222) as well as St. Michael’s Hospital (20-080) and participating POND sites including Holland-Bloorview Rehabilitation Hospital (0086), McMaster Children’s Hospital (10948), Queen’s University (6005107), and The Lawson Research Institute (115934).

## Consent to Participate

Informed consent was obtained from all individual participants included in the study.

