## Supplemental information for "The Trajectory of Depression and Anxiety Among Children and Adolescents over Two Years of the COVID-19 Pandemic"

**Supplemental Figure 1.** ***Timeline of COVID-19 Related Lockdowns and Restrictions in Ontario***

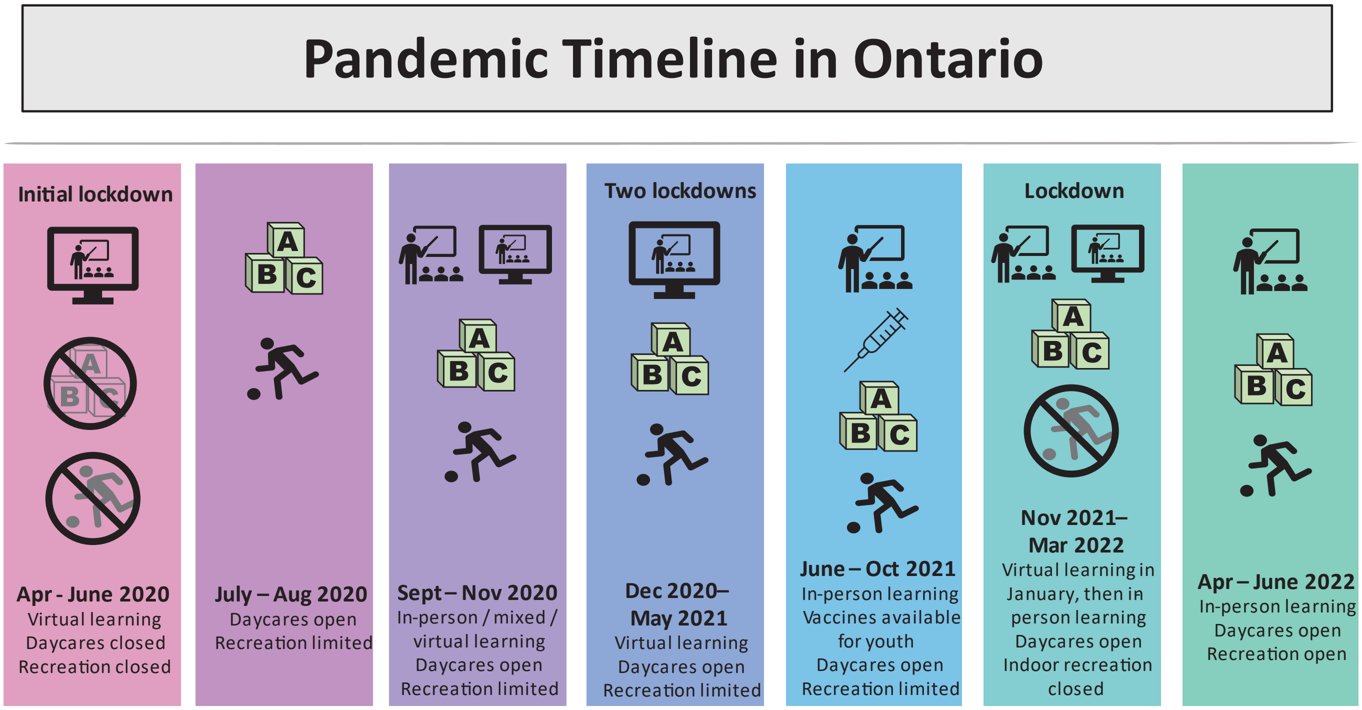

**Supplemental Figure 2.** ***Clinically Significant Depression or Anxiety Symptoms Among 8- to 12-Year-Old and 13- to 18-Year-Old Children***

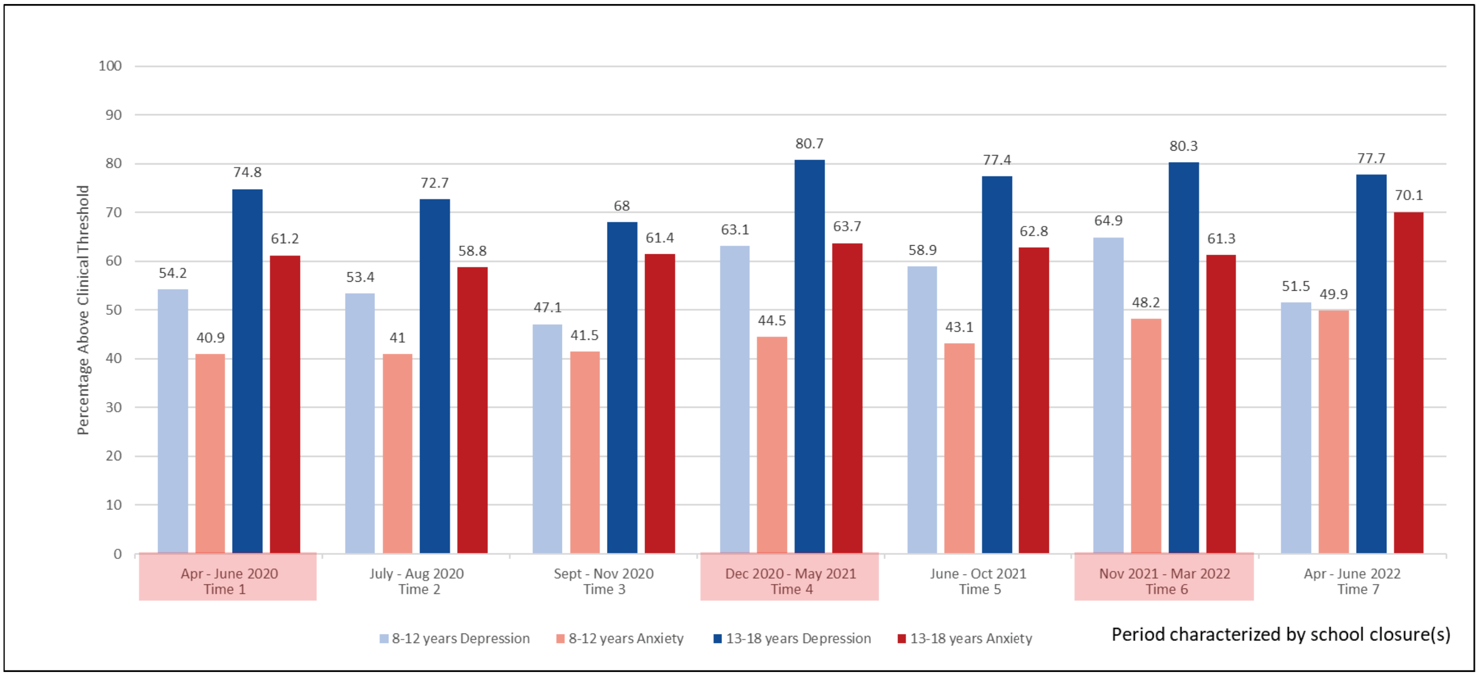

*Note.* Bar chart displays percentage of children who meet or exceed clinical threshold for Depression (RCADS or CES-DC reported by parents and children, respectively) and Anxiety (SCARED). Missing data were removed prior to visualization.

**Supplemental Table S1.** ***Characteristics of Children With and Without Pre-COVID Depression (RCADS-P) Data***

| | **With** (*n* = 330) | **Without** (*n* = 617) | Comparison |
|---|---|---|---|
| Child age – Mean (*SD*) | 11.43 (2.67) | 11.98 (2.98) | *p* = .004 |
| Assigned sex of child – % (*n*) | | | |
| Male | 51.8% (171) | 58.2% (359) | *p* = .07 |
| Female | 48.2% (159) | 41.8% (258) | *p* = .07 |
| Parent-reported Family Income – % (*n*) | | | |
| Less than $80,000 | 26.1% (86) | 21.9% (135) | *p* = .17 |
| Over $80,000 | 57.9% (191) | 63.7% (393) | *p* = .09 |
| Did not respond | 16.1% (53) | 14.4% (89) | *p* = .56 |
| Ethnicity/Ancestry – % (*n*) | | | |
| European (non-Indigenous North American) | 61.8% (204) | 68.4% (422) | *p* = .05 |
| Non-European (single ancestry origin) | 17.0% (55) | 14.4% (89) | *p* = .41 |
| Multiple ancestry origin | 20.3% (67) | 14.7% (91) | *p* = .04 |
| Did not respond | 1.2% (4) | 2.4% (15) | *p* = .30 |
| Prior Mental Health Diagnoses – % (*n*) | 67.9% (224) | 68.9% (425) | *p* = .81 |
| Pre-COVID RCADS-P Score – Mean (*SD*) | 28.01 (15.48) | .. | .. |
| Time 1 RCADS-P Score – Mean (*SD*) | 61.33 (17.33) | 62.14 (17.77) | *p* = .57 |
| Time 2 RCADS-P Score – Mean (*SD*) | 61.15 (16.88) | 62.00 (17.44) | *p* = .54 |
| Time 3 RCADS-P Score – Mean (*SD*) | 58.80 (15.77) | 60.04 (17.07) | *p* = .37 |
| Time 4 RCADS-P Score – Mean (*SD*) | 62.41 (17.85) | 63.04 (18.89) | *p* = .67 |
| Time 5 RCADS-P Score – Mean (*SD*) | 61.66 (17.05) | 61.67 (17.68) | *p* = .99 |
| Time 6 RCADS-P Score – Mean (*SD*) | 63.75 (17.55) | 63.77 (18.67) | *p* = .99 |
| Time 7 RCADS-P Score – Mean (*SD*) | 60.17 (17.24) | 58.59 (16.94) | *p* = .31 |

*Note.* RCADS-P = Revised Children's Anxiety and Depression Scale. Comparisons for age, pre-COVID RCADS-P score, and Time 1–7 RCADS-P score were computed with Welch's t-tests. All other comparisons were computed with $\chi^2$ tests of equal proportions.

**Supplemental Table S2.** ***Model Fit Indices For Conditional Models Including All Covariates***

| | $\chi^2$ | *df* | *p* | SRMR | RMSEA | CFI | TLI |
|---|---|---|---|---|---|---|---|
| Parent-report (8-18 years) | | | | | | | |
| Anxiety | | | | | | | |
| | 182.59 | 96 | < .001 | 0.04 | 0.04 | 0.98 | 0.97 |
| Depression | | | | | | | |
| | 301.14 | 96 | < .001 | 0.05 | 0.07 | 0.93 | 0.92 |
| Child-report (10-18 years) | | | | | | | |
| Anxiety | | | | | | | |
| | 150.82 | 96 | < .001 | 0.05 | 0.05 | 0.97 | 0.97 |
| Depression | | | | | | | |
| | 186.29 | 96 | < .001 | 0.06 | 0.06 | 0.95 | 0.94 |
| Parent-report (2-7 years) | | | | | | | |
| MH Problems | 71.14 | 48 | .02 | 0.07 | 0.04 | 0.97 | 0.96 |

*Note.* MH = mental health. Models satisfied cut-offs suggested by Hu & Bentler (1999): SRMR < 0.08, RMSEA < 0.08, CFI/TLI > 0.90.

**Supplemental Table S3.** ***Parameters for Growth Curve Models of Mental Health Outcomes by Parent Report Across MH Vulnerability Groups***

| | | Unconditional Model | | | | Conditional Model with Time Invariant Covariates | | | |
|---|---|---|---|---|---|---|---|---|---|
| **Outcome** | | **Mean** | | **Variance** | | **Mean** | | **Variance** | |
| **Group** | | **Estimate** | ***p*** | **Estimate** | ***p*** | **Estimate** | ***p*** | **Estimate** | ***P*** |
| **RCADS-P** | | | | | | | | | |
| **No MH** | | | | | | | | | |
| | Intercept | **52.86 (0.90)** | < .001 | **165.70 (26.85)** | < .001 | **47.03 (4.04)** | < .001 | **120.08 (19.21)** | < .001 |
| | Slope | 0.06 (0.17) | .71 | **2.60 (0.81)** | .001 | 0.11 (0.81) | .90 | **2.40 (0.78)** | .002 |
| **MH** | | | | | | | | | |
| | Intercept | **65.20 (0.68)** | < .001 | **209.08 (15.60)** | < .001 | **56.46 (3.21)** | < .001 | **181.97 (15.13)** | < .001 |
| | Slope | 0.14 (0.13) | .31 | **3.44 (0.66)** | < .001 | -0.38 (0.64) | .55 | **3.35 (0.66)** | < .001 |
| **SCARED-P** | | | | | | | | | |
| **No MH** | | | | | | | | | |
| | Intercept | **4.28 (0.25)** | < .001 | **13.21 (1.52)** | < .001 | 1.45 (1.06) | .17 | **11.03 (1.48)** | < .001 |
| | Slope | **0.19 (0.05)** | < .001 | **0.24 (0.06)** | < .001 | **0.46 (0.22)** | .04 | **0.21 (0.05)** | < .001 |
| **MH** | | | | | | | | | |
| | Intercept | **8.74 (0.21)** | < .001 | **21.42 (1.19)** | < .001 | **4.44 (0.95)** | < .001 | **18.96 (1.20)** | < .001 |
| | Slope | **0.13 (0.04)** | < .001 | **0.28 (0.06)** | < .001 | 0.16 (0.17) | .36 | **0.27 (0.06)** | < .001 |

*Note.* Estimates are in unstandardized units. Unconditional models refer to models without covariates included, whereas conditional models refer to models that adjust for covariates. Bolded estimates are statistically significant at $p < .05$. Standard error of the means are reported in parentheses. MH = prior mental health history. Depression is measured with RCADS-P (Revised Children's Anxiety and Depression Scale). Anxiety is measured with SCARED-P (Screen for Child Anxiety Related Disorders-Parent Version).

**Supplemental Table S4**. ***Parameters for Growth Curve Models of Mental Health Outcomes by Child Report Across MH Vulnerability Groups***

| | | **Unconditional Model** | | | | **Conditional Model with Time Invariant Covariates** | | | |
|---|---|---|---|---|---|---|---|---|---|
| **Outcome** | | **Mean** | | **Variance** | | **Mean** | | **Variance** | |
| **Group** | | **Estimate** | ***p*** | **Estimate** | ***p*** | **Estimate** | ***p*** | **Estimate** | ***P*** |
| **CES-DC** | | | | | | | | | |
| **No MH** | | | | | | | | | |
| | Intercept | **17.08 (1.01)** | < .001 | **151.08 (19.69)** | < .001 | -0.68 (5.33) | .90 | **116.40 (17.57)** | < .001 |
| | Slope | **0.55 (0.18)** | .003 | **3.24 (0.88)** | < .001 | -1.15 (1.02) | .26 | **3.03 (0.86)** | < .001 |
| **MH** | | | | | | | | | |
| | Intercept | **22.45 (0.73)** | < .001 | **140.06 (12.01)** | < .001 | 4.64 (4.07) | .25 | **121.70 (11.39)** | < .001 |
| | Slope | **0.32 (0.15)** | .03 | **3.39 (0.74)** | < .001 | 0.89 (0.84) | .29 | **3.24 (0.68)** | < .001 |
| **SCARED-C** | | | | | | | | | |
| **No MH** | | | | | | | | | |
| | Intercept | **6.10 (0.40)** | < .001 | **21.49 (2.81)** | < .001 | -0.44 (2.34) | .84 | **16.13 (2.65)** | < .001 |
| | Slope | **0.27 (0.08)** | < .001 | **0.42 (0.14)** | .003 | -0.40 (0.42) | .34 | **0.39 (0.13)** | .003 |
| **MH** | | | | | | | | | |
| | Intercept | **9.08 (0.30)** | < .001 | **23.62 (1.68)** | < .001 | 0.57 (1.61) | .72 | **19.63 (1.63)** | < .001 |
| | Slope | **0.14 (0.06)** | .01 | **0.53 (0.10)** | < .001 | 0.26 (0.34) | .45 | **0.52 (0.10)** | < .001 |

*Note.* Estimates are in unstandardized units. Bolded estimates are statistically significant at $p < .05$. Standard error of the means are reported in parentheses. [1]No MH group models with SCARED-C (child report) have the slope's variance fixed to 1 to improve estimation of variances. MH = prior mental health history. Depression is measured with CES-DC (Center for Epidemiological Studies Depression Scale for Children). Anxiety is measured with SCARED-C (Screen for Child Anxiety Related Disorders-Child Version).

**Supplemental Table S5.** ***Conditional Model Parameters (With All Covariates) For Children Ages 2-7***

| | **SDQ** | |
|---|---|---|
| **Time** | **Mean** | |
| **Covariates** | **Estimate (*SE*)** | ***p*** |
| **Intercept** | **7.70 (1.06)** | < .001 |
| **Variance of Intercept** | **12.84 (1.64)** | < .001 |
| Income | -0.56 (0.66) | .40 |
| Ethnicity | 0.50 (0.57) | .38 |
| Sex | **-1.77 (0.51)** | .001 |
| Age | 0.02 (0.20) | .90 |
| Parent Anxiety | **0.28 (0.06)** | < .001 |
| **Slope** | -0.12 (0.30) | .69 |
| **Variance of Slope** | **0.47 (0.18)** | .01 |
| Income | -0.29 (0.18) | .12 |
| Ethnicity | 0.05 (0.16) | .75 |
| Sex | -0.06 (0.14) | .66 |
| Age | 0.10 (0.06) | .07 |
| Parent Anxiety | -0.02 (0.02) | .36 |
| **Intercept-Slope Correlation** | -0.25 (0.48) | .61 |

*Note.* Estimates are in unstandardized units. Bolded estimates are statistically significant at $p < .05$. Standard errors of the means are reported in parentheses. MH = Prior mental health history. SDQ = Strengths and Difficulties Questionnaire.

**Supplemental Table S6.** ***Mental Health Outcomes Across Time Points by Age and Prior Mental Health Diagnoses***

| | | | Time 1 | Time 2 | Time 3 | Time 4 | Time 5 | Time 6 | Time 7 |
|---|---|---|---|---|---|---|---|---|---|
| Parent-report (8-18 years) | *n* | Measure | Mean (*SD*) | Mean (*SD*) | Mean (*SD*) | Mean (*SD*) | Mean (*SD*) | Mean (*SD*) | Mean (*SD*) |
| No MH | | | | | | | | | |
| | 293 | RCADS-P | 53·61 (15·54) | 52·57 (14·25) | 50·81 (12·74) | 54·74 (15·74) | 52·69 (13·64) | 56·11 (15·64) | 51·97 (14·33) |
| | 289 | SCARED-P | 4·51 (4·18) | 4·50 (4·29) | 4·48 (4·07) | 4·81 (4·59) | 4·75 (4·59) | 5·54 (4·76) | 5·34 (4·55) |
| MH | | | | | | | | | |
| | 635 | RCADS-P | 65·34 (17·09) | 65·34 (16·79) | 63·24 (16·61) | 67·16 (18·36) | 66·20 (17·34) | 67·92 (17·99) | 63·52 (17·15) |
| | 616 | SCARED-P | 8·77 (5·16) | 9·01 (5·17) | 8·98 (5·16) | 9·11 (5·33) | 9·03 (5·20) | 9·30 (5·28) | 9·92 (5·24) |
| Child-report (10-18 years) | | | | | | | | | |
| No MH | | | | | | | | | |
| | 186 | CES-DC | 17·18 (12·78) | 17·03 (13·23) | 16·82 (12·15) | 18·70 (13·70) | 18·85 (14·08) | 19·71 (12·92) | 20·48 (13·20) |
| | 186 | SCARED-C | 5·92 (4·69) | 6·11 (5·56) | 6·50 (5·06) | 7·20 (5·74) | 6·43 (5·65) | 7·18 (5·58) | 7·88 (6·03) |
| MH | | | | | | | | | |
| | 363 | CES-DC | 23·21 (13·07) | 22·31 (12·20) | 21·90 (13·91) | 23·47 (13·55) | 22·70 (12·58) | 25·59 (13·54) | 24·07 (12·89) |
| | 364 | SCARED-C | 9·18 (5·36) | 9·12 (5·29) | 9·65 (5·40) | 9·40 (5·40) | 9·32 (5·12) | 9·84 (5·57) | 10·22 (5·18) |
| Parent-report (2-7 years) | | | | | | | | | |
| | 327 | SDQ | 8·41 (4·28) | 8·19 (5·04) | 7·96 (4·71) | 8·09 (4·89) | 8·45 (5·24) | 7·81 (5·03) | 7·74 (5·75) |

*Note.* MH = prior mental health history. Depression is measured with RCADS (Revised Children's Anxiety and Depression Scale) and CES-DC (Center for Epidemiologic Studies Depression Scale for Children). Anxiety is measured with Parent and Child versions of SCARED (Screen for Child Anxiety Related Disorders). Mental health problems for children 2-7 years are measured using the Total Difficulties score of the SDQ (Strengths and Difficulties Questionnaire

**Supplemental Table S7.** ***Differences in Proportion of Children Above the Clinical Threshold Between Timepoints Across the Study Period***

| | T1-T7 | | T1-timepoint with most above threshold* | | T1-timepoint with least above threshold* | | T7- timepoint with most above threshold* | | T7- timepoint with least above threshold* | |
|---|---|---|---|---|---|---|---|---|---|---|
| | $\chi^2$ (*df*) | *p* | $\chi^2$ (*df*) | *p* | $\chi^2$ (*df*) | *p* | $\chi^2$ (*df*) | *p* | $\chi^2$ (*df*) | *p* |
| Depression | | | T1-T6* | | T1-T3* | | T7-T6* | | T7-T3* | |
| 8-12 | 0.82 (1) | .36 | 8.45 (1) | .004 | 4.34 (1) | .04 | 19.34 (1) | < .001 | 1.40 (1) | .24 |
| 13-18 | 0.03 (1) | .86 | 2.03 (1) | .15 | 11.26 (1) | < .001 | 0.96 (1) | .33 | 3.03 (1) | .08 |
| No MH | 2.25 (1) | .13 | 2.04 (1) | .15 | 2.89 (1) | .08 | 7.23 (1) | .01 | 0.35 (1) | .57 |
| MH | 0.17 (1) | .68 | 8.86 (1) | .003 | 11.37 (1) | < .001 | 10.87 (1) | < .001 | 4.01 (1) | .05 |
| Anxiety | | | T1-T7* except for no MH group (T1-T6) | | T1-T2* | | Only valid for no MH group (T7-T6)* | | T7-T2* | |
| 8-12 | 12.44 (1) | < .001 | … | | 2.62 (1) | .11 | … | | 11.86 (1) | .001 |
| 13-18 | 0.96 (1) | .33 | … | | 3.35 (1) | .07 | … | | 2.89 (1) | .09 |
| No MH | 4.11 (1) | .04 | 0.46 (1) | .50 | 6.26 (1) | .01 | 0.66 (1) | .42 | 7.31 (1) | .01 |
| MH | 8.17 (1) | .004 | … | | 1.92 (1) | .17 | … | | 7.22 (1) | .01 |

*Note.* MH = prior mental health history.

**Supplemental Table S8.** ***Cronbach's Alpha Reliability Coefficients for Mental Health Measures***

| Measure | Age Group | | MH Status | |
|---|---|---|---|---|
| Parent-report (8-18 years) | 8-12 years | 13-18 years | No MH | MH |
| RCADS-P | 0.86 – 0.90 | 0.88 – 0.93 | 0.82 – 0.90 | 0.85 – 0.89 |
| SCARED-P | 0.91 – 0.93 | 0.88 – 0.93 | 0.88 – 0.94 | 0.91 – 0.92 |
| Child-report (10-18 years) | 10-12 years | 13-18 years | No MH | MH |
| CES-DC | 0.90 – 0.94 | 0.91 – 0.95 | 0.88 – 0.95 | 0.92 – 0.95 |
| SCARED-C | 0.88 – 0.95 | 0.91 – 0.93 | 0.89 – 0.95 | 0.90 – 0.95 |

*Note.* Ranges of Cronbach's alpha values across all time points are listed by mental health measure, age group, or status.
